# Acute Intermittent Hypoxia Induces Motor and Cognitive Plasticity in Persons with Relapsing Remitting Multiple Sclerosis

**DOI:** 10.1101/2024.02.17.24302733

**Authors:** Milap S. Sandhu, Robert W. Motl, William Z. Rymer, Sherri L. LaVela

**Author notes:** **Corresponding Author**: Milap Sandhu, PT, PhD, 355 E Erie St Chicago, IL 60611. **Conflict of Interest**: The authors declare no competing financial interests. **Search Terms**: neuroplasticity, multiple sclerosis, acute intermittent hypoxia, rehabilitation.

## Abstract

**Background:** MS significantly impacts motor and cognitive function, yet therapies to effectively address these impairments remain limited. This study explores acute intermittent hypoxia (AIH) as a novel intervention for enhancing neuroplasticity and functional improvement in individuals with MS.

**Objective:** To examine the efficacy of a single AIH session in improving spinal motor output and cognitive performance in MS.

**Methods:** A randomized, blinded, placebo-controlled and crossover study was done in 10 individuals with relapsing-remitting MS. Participants underwent both AIH and sham AIH on separate days. AIH consisted of 15 brief exposures of low oxygen (9% O_2_) alternating with normoxia (i.e., room air). Sham AIH comprised of normoxic episodes. Pre- and post-intervention evaluations included isometric ankle torque to assess motor strength and standardized tests to evaluate cognitive function.

**Results:** Participants showed a significant increase in both plantarflexion and dorsiflexion ankle torque (p < .05), alongside significant enhancements in cognitive processing speeds as measured by the Symbol Digit Modalities Test (p < .01) after AIH. No changes were observed in auditory/verbal memory, and no adverse events were reported.

**Conclusion:** AIH presents a promising intervention for inducing neuroplasticity and improving rehabilitation outcomes in MS, suggesting the need for further exploration into its long-term impacts and mechanisms.

## Introduction

Persons with multiple sclerosis (PwMS) frequently experience movement and cognitive deficits that adversely impact their daily activities, resulting in decreased independence and quality of life.^1^ Although recent advancements in disease-modifying drugs have shown progress in altering the progression of MS and lowering relapse rates, their effectiveness in addressing established motor and cognitive impairments is still limited. The efficacy of current rehabilitative therapies in MS is also often constrained,^2^ in part due to insufficient neural activation resulting from multifocal demyelinated plaques.^3^ Thus, approaches that can augment neuroplasticity and strengthen existing neural connections represent a promising strategy. Development of novel therapeutic approaches based on the principles of neuroplasticity is important for enhancing rehabilitation outcomes, and is a major goal of MS rehabilitation research.^4^

Over the past decade, studies have shown that repeated and modest reduction in oxygen partial pressure in the inspired air (termed acute intermittent hypoxia, or AIH) can rapidly enhance neuroplasticity in individuals with incomplete spinal cord injury (SCI).^5,6^ AIH triggers the release of serotonin,^7–9^ and increases synthesis of plasticity-related proteins such as brain-derived neurotrophic factor (BDNF), resulting in rapid increases in synaptic strength and motor neuron excitability.^8–10^ This plasticity is manifested by a rapid increase in voluntary muscle strength, emerging within 60-90 minutes, in both lower- and upper-limb limbs.^11–13^ Moreover, AIH applied repetitively in combination with intensive daily rehabilitation training further enhances function, and increases both muscle strength and muscular coordination. Specifically, AIH combined with walking training is more effective in promoting locomotion speed and walking distance than either intervention delivered separately.^14,15^

While AIH has shown potential in individuals with SCI, it’s efficacy in improving function in PwMS remains undetermined. Several studies indicate a preserved potential for neuroplasticity and motor performance improvements even at higher levels of disease burden in people with MS,^16,17^ suggesting that MS patients could benefit from interventions that target functional neuroplasticity. The purpose of this study was to test the hypothesis that a single session of AIH will induce therapeutic plasticity and elicit improvement of voluntary strength and cognitive function in individuals with relapsing remitting MS.

## Methods

### Participants

All experiments were performed at the Shirley Ryan AbilityLab, an academic affiliate of Northwestern University, Chicago. The study protocol was approved by the Northwestern University (STU00207024) and the Hines Veteran Affairs (1147806) Institutional Review Boards, and informed consent was obtained from all study subjects (ClinicalTrials.gov ID = NCT04280484). Ten individuals with relapsing remitting MS and with patient-determined disease steps (PDDS) scale score of between 3 and 5 (3=gait disability, 5=need cane to walk 25 feet) participated in the study. Participants were relapse free for at least 3 months and had volitional ankle plantar flexion strength in at least one leg. Individuals with cardiovascular or respiratory disorders, obstructive sleep apnea, pregnant or nursing, or taking anti-spasticity medications were excluded.

### Study Protocol

A randomized, blinded, cross-over and placebo-controlled study design was utilized. The participants visited the laboratory twice and received either AIH or sham AIH, in a randomized order, followed by a minimum of one week of washout prior to the second visit.

Outcome measures were assessed at baseline, and 0, 30, and 60 minutes post-intervention during each visit. Study design is illustrated in Figure 1.

**Figure 1.**
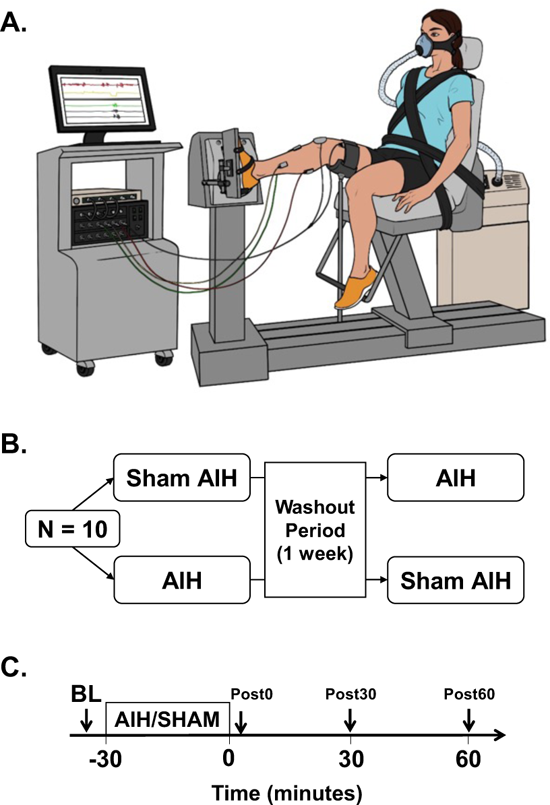
Experimental set-up. (A) Study participants were seated with the testing foot secured in a footplate coupled to a 6 degrees-of-freedom load cell. Surface electromyography was used to record muscle activity from the medial gastrocnemius, soleus, tibialis anterior and quadriceps muscles. (B) Participants received a single session of AIH or sham AIH, in a randomized order, at least one week apart. The AIH protocol consisted of 15, 60-second alternating episodes of hypoxic air (∼9% O_2_) with normoxic room air (21% O_2_). Sham AIH consisted of alternating episodes of normoxic room air. (C) Ankle plantarflexion and dorsiflexion strength, and surface electromyography was assessed prior to, and at 0-, 30-, and 60-minutes following each session. Cognitive tests were done at baseline and 60-minutes post-intervention only.

**Figure 2.**
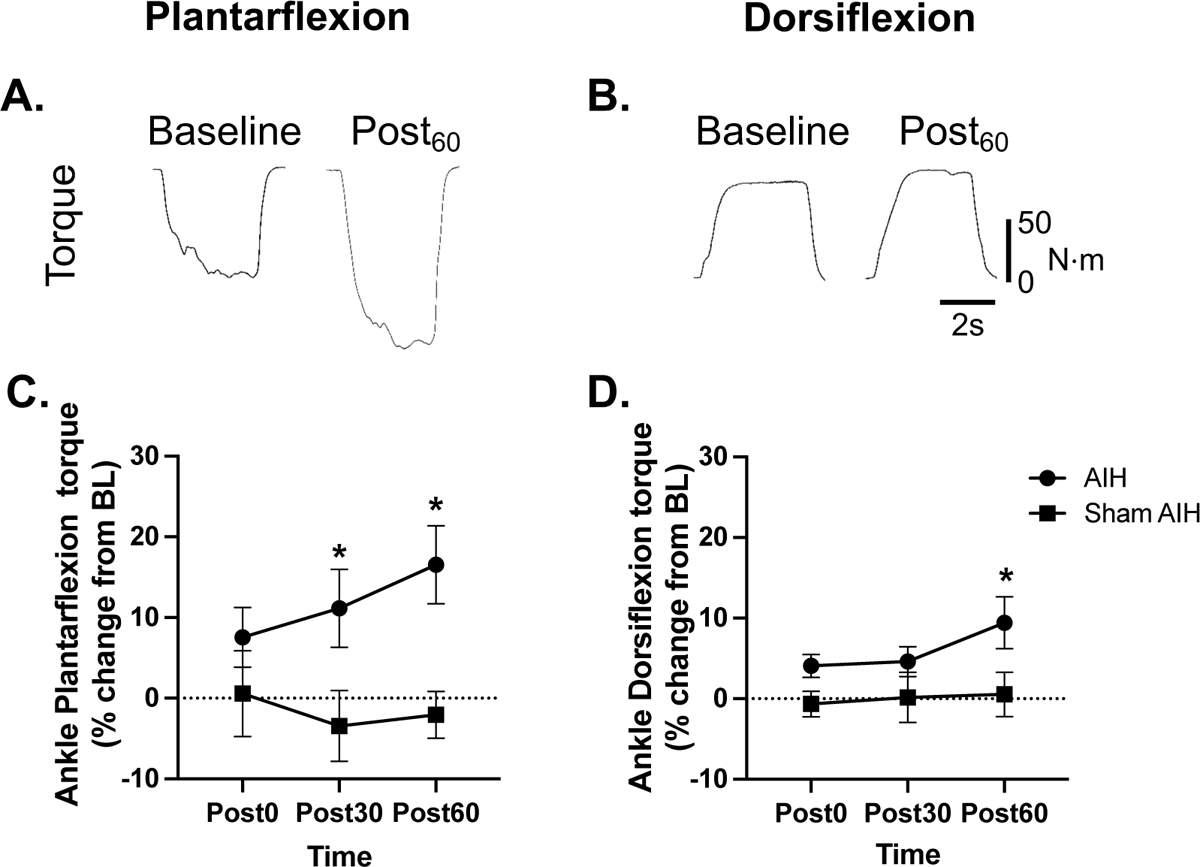
Effect of AIH on Ankle Torque. A representative example illustrating the impact of AIH on ankle plantarflexion (Panel A) and dorsiflexion (Panel B) torque. AIH or sham AIH was given immediately after BL and outcomes assessments were done at BL, immediately after intervention (Post_0_), at 30 minutes (Post_30_) and at 60 minutes (Post_60_) after the intervention.

**Figure 3.**
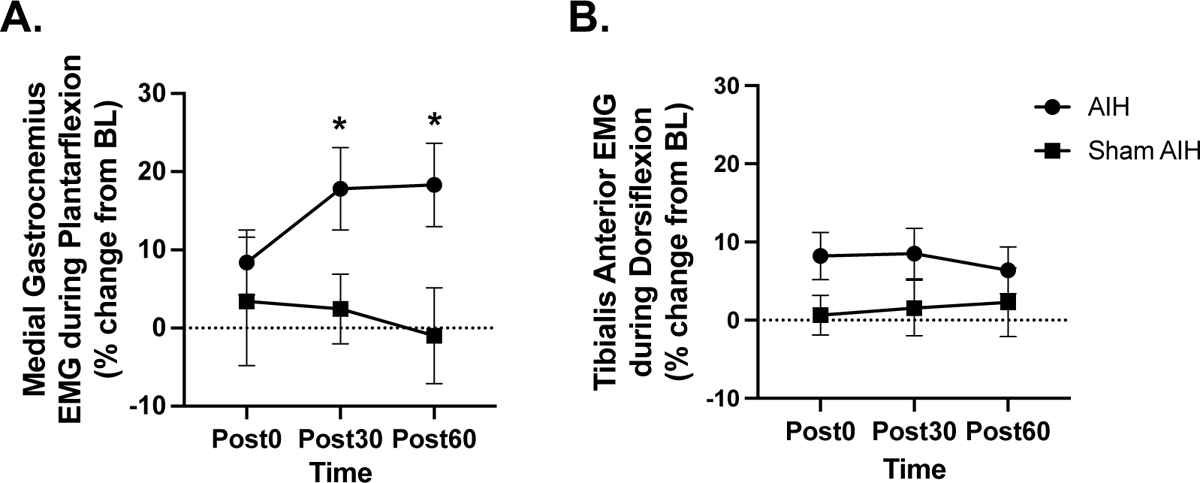
Effect of AIH on surface EMG. Mean changes in EMG activity of lower leg muscles following AIH or sham AIH are shown. Panel A shows change in medial gastrocnemius during plantarflexion and panel B shows changes in tibialis anterior during dorsiflexion. Data points represent percent change from baseline (BL). AIH or sham AIH was given immediately after BL and outcomes assessments were done at BL, immediately after intervention (Post_0_), at 30 minutes (Post_30_) and at 60 minutes (Post_60_) after the intervention. Circles denote AIH and squares denote sham AIH. *p < 0.05 vs. sham AIH.

### AIH Administration

AIH intervention was done using a hypoxia generator (Model HYP-123, Hypoxico Inc, NY, USA). Participants were fitted with a latex-free full non-rebreather mask using a custom neoprene head strap. The AIH sequence constituted 15, 60-second or less exposures of 9% O_2_ (Fraction of inspired air [FIO_2_]: 0.09), alternating with 60 seconds of 21% O_2_ (FIO_2_: 0.21). The target oxygen saturation (SpO_2_) during each hypoxia exposure was 82-88%. Sham AIH consisted of alternating exposures to normoxic air. Heart rate and SpO_2_ were recorded continuously during the AIH administration, and blood pressure measurements were done prior to and after each administration.

### Outcome Measures

Ankle joint maximum voluntary plantarflexion and dorsiflexion isometric torques were tested using a 6 degree-of-freedom load cell attached to a Biodex-4 Testing System. Subjects were secured and instructed to push or pull against the dynamometer foot plate for 3 seconds. Torque data were analog to digitally converted and sampled with EMG signals using a power 1401in the Spike2 software (CED, Cambridge, UK). Peak torques were averaged across 3 trials at each time-point, separated with 2-minute rest intervals.

Single differential surface electromyogram (EMG) electrodes were placed over medial gastrocnemius, soleus, tibialis anterior and quadriceps muscles according to the Surface ElectroMyoGraphy for the Non-Invasive Assessment of Muscles (SENIAM) guidelines. EMG data was collected with the Bagnoli EMG system, sampled at 2000 Hz, synchronized with torque data, amplified 1000 times, and filtered (bandwidth 20-450 Hz). EMG data was integrated, rectified, and smoothed offline in Spike2 software. Peak EMG activity was computed from a 100 msec time window, 150 msec preceding the maximum absolute value of the torque signal for each contraction. Average peak EMG activity for each muscle was obtained across the 3 MVCs.

Cognitive information processing speed was assessed with the Symbol Digit Modalities Test (SDMT). Participants matched symbols to numbers within 90 seconds; the maximum score is 110. Verbal learning and memory were measured using the California verbal learning test-II (CVLT-II). Participants recalled words from a read list over five trials; the highest score is 80.

### Statistical Analysis

The relative change from baseline to subsequent time points was calculated for each participant during each treatment period. The primary outcome was relative change in torque from baseline to 60 minutes post-intervention. Data were tested for normality using Shapiro-Wilk test. A mixed model was fitted using Restricted Maximum Likelihood in GraphPad Prism 9.0, and Tukey’s Honest Significant Difference test was used for multiple pairwise-comparison. A Geisser–Greenhouse correction was used to correct for lack of sphericity. Fixed effects were time, treatment (i.e. AIH or Sham AIH), and treatment interaction with time. Random effect was subjects. Data are presented as mean ± standard error (SE).

## Results

Eleven participants were enrolled in the study. One individual dropped after the first session, and their data was excluded from analysis. The remaining participants completed both sessions with no reported adverse events. Demographics of study participants are presented in Table 1.

**Table 1.** Demographic characteristics of study participants. W, white; AA, African American; BMI, body mass index; PDDS, patient-determined disease steps; treatment 0-sham AIH, 1-AIH.

| Study ID | Sex | Age Range (yr) | Race | BMI | MS Duration (yr) | PDDS | Treatment Order |
| --- | --- | --- | --- | --- | --- | --- | --- |
| 01 | M | 61-65 | W | 32.0 | 35 | 3 | 1,0 |
| 02 | M | 51-55 | AA | 26.3 | 12 | 3 | 1,0 |
| 03 | M | 61-65 | W | 22.8 | 3 | 3 | 1,0 |
| 04 | M | 56-60 | AA | 33.0 | 26 | 3 | 0,1 |
| 05 | F | 61-65 | AA | 30.2 | 16 | 5 | 0,1 |
| 07 | M | 66-70 | W | 31.9 | 16 | 3 | 0,1 |
| 08 | M | 41-45 | AA | 24.4 | 13 | 5 | 1,0 |
| 09 | M | 66-70 | AA | 20.0 | 27 | 5 | 0,1 |
| 10 | M | 71-75 | W | 29.3 | 49 | 5 | 1,0 |
| 11 | M | 51-55 | W | 28.2 | 16 | 3 | 0,1 |

### Ankle plantarflexion torque

There were no significant differences in baseline values of ankle plantarflexion strength prior to the AIH and sham AIH intervention (Pre-AIH: 67.9 ± 10.2 Nm; Pre-sham AIH: 65.3 ± 10.6 Nm, p = 0.86). Plantarflexion torque increased by 7.5 ± 3.7%, 11.1 ± 4.8% and 16.5 ± 4.8% at 0, 30 and 60 minutes after AIH, respectively. Equivalent values after the sham AIH condition were 0.6 ± 5.3%, −3.4 ± 4.4% and −0.6% ± 2.8% of baseline, respectively. A two-way repeated measures ANOVA revealed that there was a statistically significant interaction between the effects of treatment and time (F(3, 54) = 4.318, p = 0.0084). Simple main effects analysis showed that AIH treatment had a statistically significant effect on ankle strength (p = 0.026). Tukey’s multiple comparisons test revealed significantly higher torque at 60 minutes post-AIH, compared to baseline (p < 0.032). No differences were observed at any time points after sham AIH (all p > 0.83).

### Ankle dorsiflexion torque

Baseline ankle dorsiflexion torque values were not statistically different prior to AIH and sham AIH (AIH: 45.0 ± 9.7 Nm; sham AIH: 41.7 ± 8.8 Nm, p = 0.8). Post-AIH torque was 4.1 ± 1.4%, 4.6 ± 1.9% and 9.4 ± 3.1% higher than baseline at 0, 30 and 60 minutes after AIH. Equivalent values for the sham AIH condition were −0.7 ± 1.6%, 0.2 ± 3.1% and 0.5% ± 2.7%, respectively. A two-way repeated measures ANOVA revealed that there was no statistically significant interaction between the effects of treatment and time (F(3, 48) = 2.212, p = 0.0987). Simple main effects analysis showed that treatment had a statistically significant effect on ankle strength (p = 0.0476). Tukey’s multiple comparisons test revealed significantly higher torque at 30 and 60 minutes post-AIH, compared to baseline (both p ≤ 0.0487). No differences were observed at any time points after sham AIH (all p > 0.952).

Mean changes in plantarflexion and dorsiflexion torque after a single session of AIH or sham AIH are presented in Panels C and D, respectively. Data points represent percent change from baseline (BL). Circles denote AIH and squares denote sham AIH. *p < 0.05 vs. sham AIH.

### EMG Response

There was a significant increase in medial gastrocnemius EMG activity after AIH. Compared to baseline, medial gastrocnemius EMG was 8.3 ± 4.2%, 17.8 ± 5.3% and 18.3 ± 5.0% at 0, 30 and 60 minutes post-AIH, respectively. Simple main effects analysis showed that treatment had a statistically significant effect on ankle strength (p = 0.0385). Tukey’s multiple comparisons test revealed significantly higher torque at 30 and 60 minutes post-AIH, compared to baseline (both p ≤ 0.0453). There were no significant effects of AIH on the EMG activity of other muscles, and sham AIH on the EMG of any muscle. However, there was a notable trend towards significance in TA activity during dorsiflexion at Post_0_ after AIH, with a p-value of 0.07 compared to sham AIH.

### Cognitive function

There were no significant differences in baseline scores of SDMT prior to the AIH and sham AIH intervention (Pre-AIH: 37.8 ± 2; Pre-sham AIH: 42.3 ± 3.9, p = 0.21). SDMT scores increased by 27 ± 7% at 60 minutes after AIH (37.8 ± 2 to 47.9 ± 3.4). Equivalent values after the sham AIH condition were −5 ± 5% (42 ± 3.5 to 40 ± 3). Mixed-effects model analysis indicated a statistically significant main effect of time on SDMT performance (F (1, 16) = 6.758, p = 0.0194). In contrast, the main effect of treatment was not significant (F (1, 16) = 0.1184, p = 0.7352). However, the interaction between time and treatment was significant (F (1, 16) = 15.07, p = 0.0013), suggesting differential effects of time on SDMT performance based on treatment type. Bonferroni’s multiple comparisons test revealed significantly higher SDMT post-AIH, compared to baseline (p < 0.05). There were no significant changes in CVLT-II scores following AIH or sham AIH.

## Discussion

This study provides evidence that intermittent hypoxia protocols can elicit motor and cognitive changes in patients with relapsing-remitting MS. In our experiments, voluntary ankle plantarflexion and dorsiflexion strength increased after a single session of AIH. We also found that memory processing speed, as measured with SDMT, improved in our study participants. Most importantly, AIH was well tolerated by participants and no adverse events were reported, suggesting its feasibility as a therapeutic modality in PwMS.

In the past decade, several studies have shown that AIH can rapidly induce neuroplasticity in neurologically intact individuals as well as persons with SCI.^11,12,18–20^ This neuroplasticity manifests as enhanced volitional somatic motor output in the upper and lower-limbs, generating 20-40% increases in force within 60-90 minutes. Our study builds on the existing evidence of AIH-induced plasticity in SCI population and extend these findings to another clinical condition that often limits motor function. Prior work suggests a role of corticospinal pathways as evidenced by an increase in motor evoked potentials following AIH. AIH has been shown to enhance synaptic strength within descending neural pathways in spinal motor neurons and enhance corticospinal excitability in individuals with SCI.^21^ Whether AIH exerts its effects through a similar mechanism in MS is not known. The magnitude of change in ankle strength after AIH in MS is similar to that seen in SCI population,^12,22^ suggesting that similar mechanisms to those elucidated in SCI underlie the effects reported here.

Recent work by Tokarska and colleagues presents alternate potential AIH mechanism in MS.^23^ In this study, experimental autoimmune encephalomyelitis (EAE) mice, a model for MS, were subjected to daily AIH (10 cycles of 5-min each) for 7 days, beginning at near peak EAE disease score. They demonstrated that AIH not only altered the disease course but also promoted intrinsic repair and functional recovery. Importantly, their study revealed that AIH treatment led to enhancement of myelination, axon protection, and oligodendrocyte precursor cell recruitment to demyelinated areas. They also found a dramatic reduction in inflammation, with remaining macrophages/microglia polarized toward a pro-repair state. This is particularly significant given the extensive demyelination characteristic of MS. Although a single session of AIH is unlikely to affect inflammation or disease course, this is an important observation in context of future studies using repetitive AIH.

An important observation in our results was the differential response in torque changes between plantarflexion and dorsiflexion following AIH. Specifically, plantarflexion torque demonstrated an increase of 16.5 ± 4.8% at 60 minutes post-AIH, which was markedly higher than the dorsiflexion torque change of 9.4 ± 3.1% during the same time interval. This distinction may reflect distinct underlying neural control mechanisms and brain activation patterns during these two movements. fMRI studiers shows that dorsiflexion engages more extensive cortical activation, including the left medial M1 and bilateral supplementary motor areas, and plantarflexion predominantly involves subcortical areas, suggesting different demands on cortical resources.^24,25^ The more significant change in plantarflexion torque following AIH might be due to a more considerable plasticity potential or responsiveness of the subcortical areas compared to the cortical structures that are more involved with dorsiflexion. Further research, particularly using transcranial magnetic stimulation techniques to probe cortical vs subcortical areas could further elucidate the distinct neural substrates involved in AIH-induced plasticity.

One of the intriguing findings in our study was the marked improvement in cognitive processing following AIH treatment, as indicated by the notable increase in SDMT scores. The increase of 10.1 points from pre to post-AIH SDMT scores is significantly higher than the clinically meaningful threshold of 4 points.^26^ The relationship between AIH and BDNF expression could offer a potential explanation for this finding. Animal studies suggest that AIH upregulates BDNF protein levels in the CNS and facilitates BDNF-dependent signaling cascades.^8,9,27,28^ Considering that epigenetic-mediated changes in BDNF gene expression impact cognitive processing also,^29^ the increase in BDNF levels induced by AIH could be responsible for the cognitive improvements we observed. Additional larger studies are warranted to confirm this hypothesis.

### Limitations

A constraint of this study is a relatively higher number of males than females. Our study recruitment was limited to the local Veteran Affairs hospital, where the patients are predominantly male veterans. Given that sex differences may substantially influence the response to AIH through disparate hormonal interactions with neuroplastic processes,^30^ future studies should aim for a more balanced sex representation and target general demographics in MS, which is more prevalent in women. A second limitation of our study is that the exploratory nature of our project limited our capacity to identify the underlying factors that may contribute to the variability in AIH responsiveness among PwMS. Future studies powered to detect covariates of responsiveness, such as sex, race and baseline disability, are needed.

## Conclusions

This study demonstrates that AIH is as a promising intervention for enhancing motor and cognitive function in PwMS. The observed improvements, along with no adverse events, highlight the feasibility and efficacy of this approach in promoting neuroplasticity. These findings encourage further research into underlying mechanisms, effect of repetitive sessions, and factors influencing responsiveness in PwMS.

## Author Contributions

**Milap Sandhu:** Conceptualization, Methodology, Investigation, Formal analysis, Visualization, Writing - Original Draft preparation. **Robert Motl:** Conceptualization, Writing - Review & Editing. **William Z Rymer:** Conceptualization, Writing - Review & Editing. **Sherri LaVela:** Conceptualization, Methodology, Investigation, Formal analysis, Writing - Review & Editing, Funding acquisition.

## Data Availability

All data produced in the present study are available upon reasonable request to the authors.

